# Personalized Brain-Computer Interface-based Intervention for Mindful Anxiety Regulation in Young Adults: A Randomized Clinical Trial

**DOI:** 10.1101/2024.03.01.24303187

**Authors:** Wan Jie Tan, Yi Ding, Xiangting Bernice Lin, Neethu Robinson, Qiuhao Zeng, Su Zhang, Aung Aung Phyo Wai, Tih-Shih Lee, Cuntai Guan

## Abstract

**Importance:** Brain-computer interface-(BCI-) based interventions are promising means for self-administered anxiety treatment due to their non-invasiveness and portability. However, few studies have explored their viability in the home setting.

**Objective:** To evaluate the feasibility and preliminary efficacy of a novel, personalized, BCI-based intervention integrating mindfulness principles for anxiety regulation in the home setting.

**Design:** An open-label, two-arm randomized clinical trial was conducted from January 2021 to December 2021.

**Setting:** The study was conducted at Duke-NUS Medical School, Singapore.

**Participants:** Thirty young adults from the community aged 21-35 with at least moderate anxiety were randomized 1:1 to either the intervention or waitlist-control group.

**Intervention:** The intervention was self-administered by the participants and involved eight 30-minute sessions held at home over 2 weeks.

**Main Outcomes and Measures:** The primary outcomes were safety, acceptability, and anxiety (Beck Anxiety Inventory II, State-Trait Anxiety Inventory [Form Y], and electroencephalography [EEG] data). Secondary outcomes were quality of sleep (Insomnia Severity Index), emotion regulation (Difficulties in Emotion Regulation Scale), mindful awareness (Mindfulness Awareness Attention Scale), and depression and stress subscales in the Depression Anxiety Stress Scale-21.

**Results:** The intervention was safe and acceptable. No severe adverse events were reported. The attrition rate was 40%, predominantly due to technical issues. Pooled analyses indicated a significant reduction in anxiety, depression, stress, and insomnia from pre- to post-intervention. Improvements in anxiety were supported by physiological changes. Participants also reported significantly greater mindful awareness, access to emotion regulation strategies, and control over impulses.

**Conclusion and Relevance:** These findings suggest that our BCI-based intervention is feasible and potentially efficacious for participants to entrain anxiety regulation independently at home. A larger trial is warranted.

**Trial Registration:** ClinicalTrials.gov NCT04626713; https://tinyurl.com/27hn2m76

## Introduction

Anxiety disorders are common and characterized by excessive worry, hyper-arousal, and fear [1]. A nationwide epidemiological survey in Singapore showed that the lifetime prevalence of Generalized Anxiety Disorder rose from 0.9% in 2010 to 1.6% in 2016 [2], with young adults being particularly affected [3]. Anxiety disorders impede various domains of functioning [4, 5], yet current treatments remain suboptimal [11, 13, 14]. Thus, alternative interventions are necessary.

Mindful emotion regulation presents one promising strategy by cultivating a changing relationship with one’s anxiety [6]. This strategy facilitates reduced reactivity to emotional stimuli by encouraging one to accept and detach from their aversive feelings, thoughts, and ineffective habitual responses [7, 8]. Benefits of mindfulness include reduced anxiety and depression, and improved sleep quality, all of which lead to increased overall well-being [9, 10]. Unfortunately, individuals with no experience in such practices can find it challenging to engage in mindful emotion regulation consistently. As sustained practice is crucial to yield the benefits of mindfulness practice [11], an appealing mode of treatment delivery is necessary.

Brain-computer interface (BCI) could serve as a useful treatment modality. BCI is a specific form of biofeedback that enables the direct communication between a human brain and an external device [12]. Recent preliminary studies suggest that biofeedback-based relaxation and mindfulness training are useful for both healthy and anxious individuals [13]. Given its non-invasiveness and portability [16], and that 96–97% of individuals aged 15–34 years in Singapore regularly use technology [14], BCI technology may serve as a feasible means for self-administration of treatment at home.

Our study extends beyond laboratory trials to evaluate the feasibility and potential efficacy of a novel personalized BCI-based intervention for entraining mindful anxiety regulation at home. Mindfulness principles were integrated into a BCI-based game interface driven by a novel EEG algorithm. Thus, the present study aimed to evaluate: (1) the safety and acceptability of this intervention, and (2) its preliminary efficacy in reducing anxiety among community-dwelling young adults.

## Methods

### Trial design, Study Approval and Registration

This study is an open-label, two-arm randomized controlled feasibility trial, comparing a home-based BCI-based intervention group (INTG) to a waitlist control group (WCG). Power calculations were based on 80% safety and acceptance rate. Assuming an attrition rate of 20%, a total sample size of 30 can give a precision level (width of 95% confidence interval) of about ±20% margin of error. A sample size of 30 participants was considered adequate to find significant differences in the main outcome variables [e.g., 15, 16, 17].

Ethics approval was obtained from the Institutional Review Board of the National University of Singapore (reference code: NUS-IRB-2020-220). The study was prospectively registered on ClinicalTrials.gov prior to participant enrollment (NCT04626713). Informed consent was provided before randomization.

### Participants

Thirty young adults aged 21–35 with at least moderate anxiety were recruited from the community. To be eligible for the study, participants needed to (1) have a score of 16 and above on the Beck Anxiety Inventory II (BAI-II), (2) be computer literate, and (3) have access to a Windows 10 computer. The exclusion criteria were: (1) diagnosis of any anxiety disorder induced by a substance or medical condition; obsessive compulsive disorder; bipolar disorder; any psychotic disorder (lifetime); intellectual disability; autism spectrum disorder; attention-deficit/ hyperactivity disorder, (2) history of substance use disorder, (3) neurological disorders (e.g., epilepsy, cerebrovascular accidents), (4) metal in the cranium, skull defects, or skin lesions on scalp at proposed electrode sites, (5) severe visual or hearing impairment, or (6) prior experience with mindfulness-based therapy. Figure 1 (CONSORT flowchart) shows the participant flow throughout the study.

**Figure 1.**
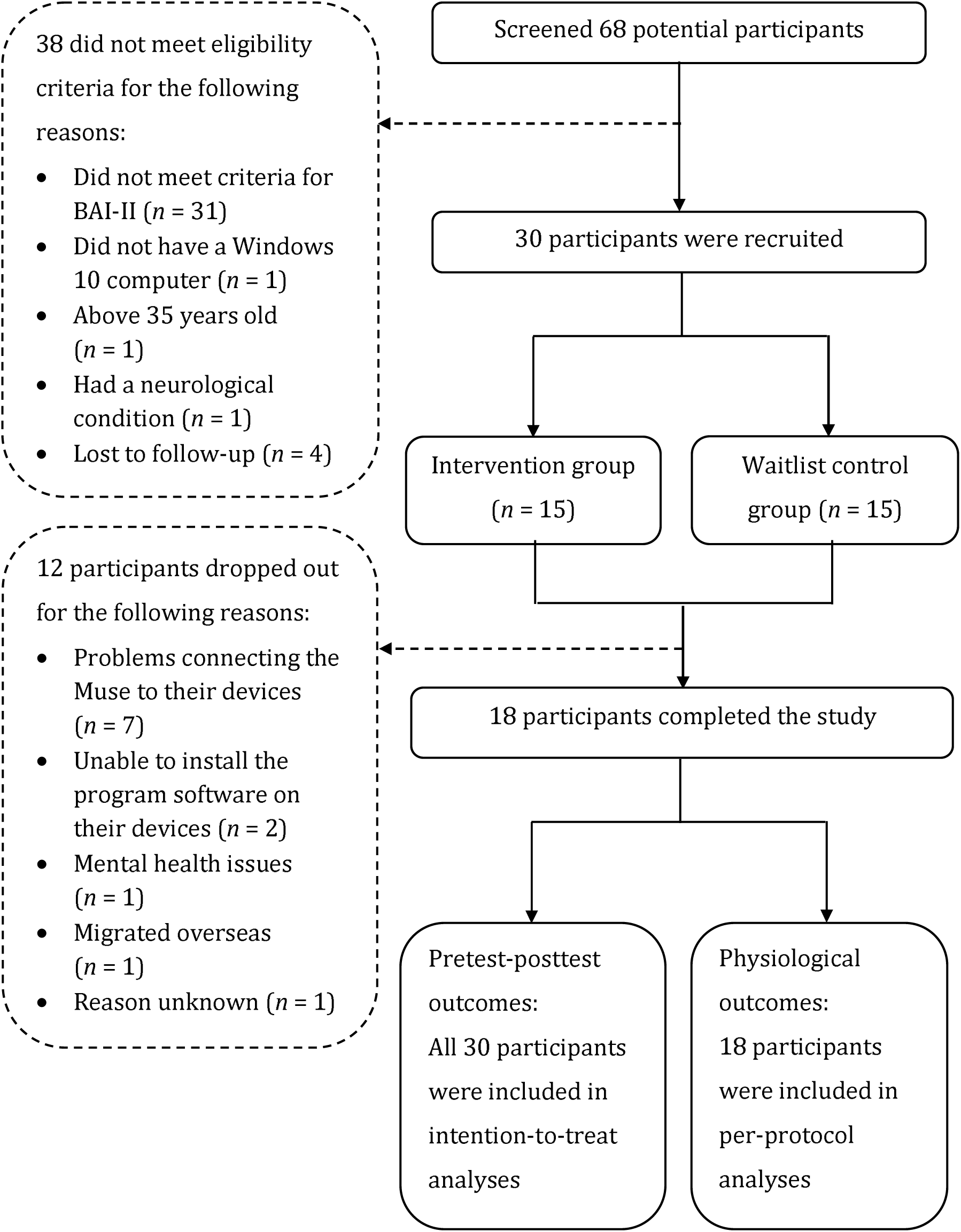
Flow chart of the study design. 30 participants were recruited and randomized into the INTG or WCG. The overall attrition rate was 40%.

### Randomization and Allocation

Participants who met the eligibility criteria were allocated to either the INTG or WCG. Randomization sequence was generated using block randomization methods in Microsoft Excel by an independent statistician who had no contact with the participants. The allocation ratio for the INTG and WCG was 1:1. Due to the study design, participants and research staff were not blinded to the treatment allocation. Analyses of physiological data were performed by personnel blinded to treatment allocation.

### Intervention Program

The intervention consists of eight 30-minutes sessions held online at home, four times a week over two weeks. Each session began with a brief interactive psychoeducation session on anxiety with an audio-guided mindfulness practice. Participants then played a 15-minute BCI-based game integrating mindfulness principles. During the game, participants wore a mobile, non-invasive EEG-biosensor device that is commercially available from Muse (Muse version 2016; InterAxon Inc., Toronto, ON, Canada).

### Study Procedure

Participants were recruited from the community through posters, social media, institutional email notices, and word-of-mouth. Recruitment was conducted on a rolling basis until a sample size of 30 was reached. Informed consent was obtained prior to screening and recruitment. Eligible participants were randomized into either the INTG or WCG. All participants visited Duke-NUS Medical School, Singapore at Week 1 for calibration and to complete a baseline assessment battery. The assessment battery comprises all outcome measures unless otherwise specified. The INTG was provided with a Muse headset and self-administered treatment sessions in Weeks 2 and 3 while the WCG continued with their usual activities. All participants returned to the study site in Week 4 to complete another assessment battery. The WCG was then provided with the Muse headset and self-administered the same sessions as the INTG in Weeks 5 and 6. Both INTG and WCG completed the assessment battery in Week 7. Reimbursement was provided on a prorated basis upon completion or termination of the study. The study was conducted from January 2021 to December 2021.

### Measures

#### Primary outcomes

Anxiety was measured using the BAI-II [18] and the State-Trait Anxiety Inventory (STAI Form Y) [19]. The BAI-II is a self-report clinical measure that assesses the severity of anxiety. It has 21 items scored on a scale of 0 (“not at all”) to 3 (“severely”). Higher scores indicate higher levels of anxiety, which can be interpreted as: 0-7 (minimal anxiety), 8-15 (mild anxiety), 16-25 (moderate anxiety), and 26 or above (severe anxiety). The STAI is a widely used self-report measure for state and trait anxiety. It consists of 20 items for each of two subscales, using a four-point Likert scale, from 1 (“not at all”) to 4 (“very much so”) for the trait anxiety factor, and from 1 (“almost never”) to 4 (“almost always”) for the state anxiety factor. Scores range between 20 and 80 for each subscale, with higher scores indicating greater anxiety. Both scales demonstrated excellent internal consistency in the present study (BAI-II: α=.90; STAI: α=.91).

A safety monitoring log was used to measure the number of adverse events reported. The total number and severity rating of all adverse events reported were collated at the end of the study. Qualitative feedback was obtained at post-intervention using questions adapted from the System Usability Scale (SUS) [20].

#### Secondary outcomes

Subjective sleep quality was measured using the Insomnia Severity Index (ISI) [21]. The ISI is a 7-item instrument assessing the severity and impact of both nighttime and daytime components of insomnia. Items are rated on a 5-point Likert scale (‘0’=not at all, ‘4’=extremely) and the total score ranges from 0 to 28. A higher score suggests more severe insomnia. The scale demonstrated good internal consistency in the present study (α=.85).

The short-form version of the Difficulties in Emotion Regulation Scale (DERS-SF) [22, 23] is an 18-item measure of emotion regulation issues. There are six sub-scales, each with three items: Strategies, Non-acceptance, Impulse, Goals, Awareness, and Clarity. Items are scored on a 5-point Likert scale ranging from 1 (“almost never”) to 5 (“almost always”). Subscale scores are obtained by summing up the corresponding items. The DERS-SF had good internal consistency in the present study (α=.83).

Mindful awareness was assessed using the Mindfulness Awareness Attention Scale (MAAS) [24]. It is a 15-item questionnaire rated on a scale from one (“almost always”) to six (“almost never”), with higher total scores indicating greater mindfulness. In the present study, the MAAS had good internal consistency (α=.83).

The 21-item version of the Depression Anxiety Stress Scale (DASS-21) [25] was used to measure the severity of symptoms of depression, anxiety and stress. There are seven items for each of the three subscales, which comprises a statement and is scored from 0 (“Did not apply to me at all”) to 3 (“Applied to me very much, or most of the time”). In the present study, the DASS-21 demonstrated excellent internal consistency (α=.92).

### BCI-based Game

The BCI-based game was developed by the research team using Unity3D. EEG data was recorded simultaneously while participants played the game. The game includes two alternating scenes: an anxiety elevation game to avoid rocks shown in Figure 2 (A) and relaxation scenes shown in Figure 1. The block diagram of the system was shown in Figure 2 (C). In the game, a rock rushes towards the participant on the screen at random time intervals. Participants were required to avoid the rock by clicking the correct key on the keyboard indicated by an arrow on the screen. In the relaxation scene, a bird flies slowly in a winter landscape, accompanied by a mindfulness-based audio guide and soft ambient music. The game entrains mindful anxiety regulation over time by alternating regularly between anxiety elevation and mindful relaxation scenes.

**Figure 2.**
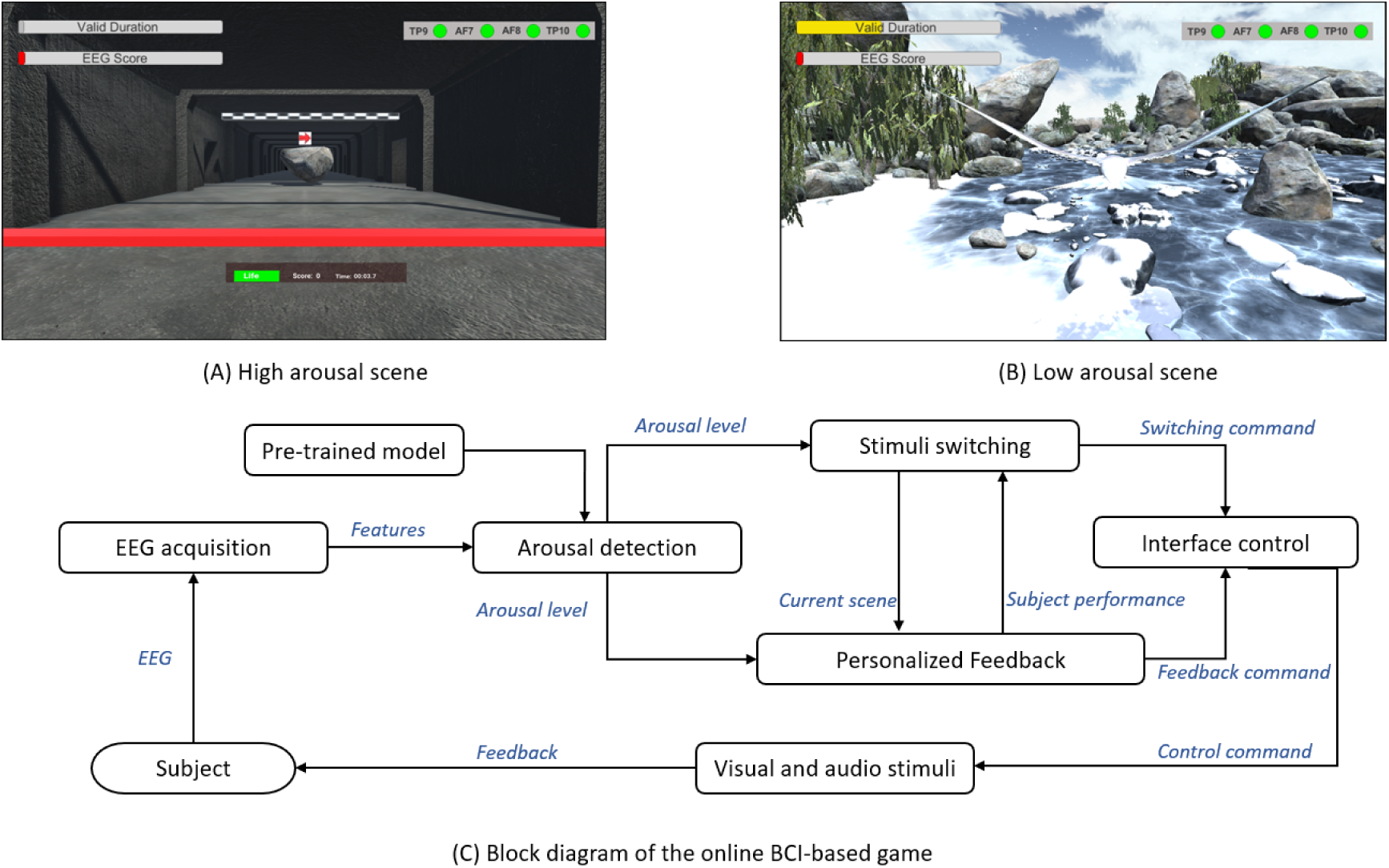
Illustration of the BCI-based game. (A) shows the high arousal scene. (B) demonstrates the low arousal scene. The block diagram is shown in (C).

All participants went through a calibration stage in Week 1, where 6-minutes of EEG data were collected to build a machine learning classifier for an online inference of the arousal level. This allows subsequent gameplay to be personalized based on individual baseline anxiety levels. Relative power features of the EEG data in different frequency bands were utilized and a Support Vector Machine (SVM) classifier was trained upon them as the personalized classifier for the participant. The trained classifier, which provides predictions of arousal levels in real-time, is then used during gameplay. Arousal levels were reflected to participants in real-time by the speed of bird wing flapping during mindful relaxation. Mindfulness-based audio guidance was triggered to play whenever the detected arousal change exceeds a stipulated threshold.

### EEG Recording

EEG signals from two frontal electrodes (AF7 and AF8) and two temporal electrodes (TP9 and TP10) were recorded. Fpz (center of the forehead) was used as reference electrodes. Data from AF7 and AF8 were utilized for EEG analyses. The sampling rate was 256 Hz. EEG data recorded in the first and the last sessions were used in pre- and post-intervention analyses.

### Data Analyses

SPSS Statistics version 27 and Python 3.10 (SciPy version 1.9.3) were used to analyse pretest-posttest self-rated outcomes and physiological outcomes, respectively. Given the small sample size, pre-and post-intervention analyses using Wilcoxon signed-rank tests were performed for each of the outcome measures (α=.05).

Distributions of the short-epoch EEG were used to capture time-based changes. Anxiety is associated with decreased slow wave brain activity (alpha) and increased fast wave brain activity (beta) [26]. This pattern of brain activity is inversed during mindfulness practices, consistent with a physiological reduction of anxiety [27]. Accordingly, EEG data were pre-processed and the Alpha (8-12 Hz), Beta-1 (12.5-16 Hz), Beta-2 (16.5-20 Hz), and Beta-3 (20.5-28 Hz) bands were derived.

Differences in EEG alpha- and beta-band power feature distributions between pre- and post-intervention were analyzed. Kolmogorov–Smirnov test was applied to test the differences between pre- and post-intervention EEG power feature distributions for each participant. Pearson correlation coefficient *r* was used to evaluate the correlations between changes in primary outcomes and mean alpha- and beta-band power features. Effect sizes were interpreted using Cohen’s *r* (0.1 as small, 0.3 as medium, and 0.5 as large) [28].

## Results

### Participants

A total of 68 potential participants from the community completed the screening questionnaire. Thirty-eight were excluded as they either did not meet the inclusion criteria or were lost to follow-up. The demographic characteristics of the remaining 30 participants are summarized in Table 1. There were no statistical differences between INTG and WCG for all demographic characteristics and baseline anxiety (*p*>.05 in all cases). The attrition rate was 40% (*n*=12). Reasons for dropping out are detailed in Figure 1. Participants who dropped out were more likely to be university graduates, *X*^2^(1)=4.00, *p*=.046.

**Table 1.** Participant and baseline characteristics.

| Characteristics | Condition |  |
| --- | --- | --- |
|  | INTG (*n* = 15) | WCG (*n* = 15) |
| Age in years, mean (*SD*) | 25.7 (2.7) | 26.27 (2.8) |
| Gender, *n* (%) |  |  |
| Male | 4 (27) | 7 (47) |
| Female | 11 (73) | 8 (53) |
| Ethnicity, *n* (%) |  |  |
| Chinese | 11 (73) | 13 (87) |
| Malay | - | 1 (7) |
| Indian | 4 (27) | - |
| Others | - | 1 (67) |
| Education, *n* (%) |  |  |
| University | 12 (80) | 13 (87) |
| Pre-university | 3 (20) | 2 (13) |
| BAI-II, mean (*SD*) | 22.2 (8.3) | 24.46 (12.8) |

*Note.* INTG = intervention group; WCG = waitlist control group; University = Bachelor's Degree or higher; Pre-university = Diploma, National ITE Certificate, or General Certificate of Education 'A'-Level

### Safety and acceptability

No adverse events were reported. Qualitative feedback suggests that participants enjoyed the psychoeducation aspect of the intervention as it was informative and interesting (6/18, 33.3%), and found the intervention helpful in guiding them to relax (6/18, 33.3%). However, participants encountered occasional disconnection from the Muse headset and were unable to play the game smoothly (6/18, 33.3%). Other suggestions included improving or varying visuals (e.g., different scenarios such as spring or summer in a field, bigger arrows; 5/18, 27.8%). One participant felt “a little depressed when looking at the snow and enclosed lake” in the game.

### Preliminary efficacy

Pre- and post-intervention outcomes were analyzed by collapsing INTG W1 data with WCG W4 data to form pre-intervention scores, and INTG W4 data with WCG W7 data to form post-intervention scores. Analyses revealed that anxiety decreased significantly across the BAI-II, STAI-T, and STAI-S from pre- to post-intervention (Table 2). Significant improvements were also found on the ISI, MAAS, strategies and impulse measured on the DERS-SF, and all subscales of the DASS-21. These analyses demonstrated medium to large effect sizes.

**Table 2.** Pre- and post-intervention analyses for pooled data.

|  | INTG median (IQR) |  |  | WCG median (IQR) |  |  | Pooled data median (IQR) |  | *z* | *p* | Effect size (*r*) |
| --- | --- | --- | --- | --- | --- | --- | --- | --- | --- | --- | --- |
|  | Week 1 | Week 4 | Week 7 | Week 1 | Week 4 | Week 7 | Pre-intervention | Post-intervention |  |  |  |
| Primary outcome measures |  |  |  |  |  |  |  |  |  |  |  |
| BAI-II | 20.0<br>(17.0-24.0) | 16.7<br>(11.5-16.7) | 11.9<br>(10.9-11.9) | 24.5<br>(18.5-29.0) | 25.2<br>(21.0-30.0) | 19.2<br>(18.0-22) | 23.6<br>(18.3-29.3) | 18.0<br>(16.0-20.5) | 4.08 | <.001 | 0.74 |
| STAI |  |  |  |  |  |  |  |  |  |  |  |
| State anxiety | 50.6<br>(48.0-54.5) | 40.9<br>(38.0-42.9) | 42.1<br>(42.1-45.5) | 48.0<br>(43.5-56.0) | 46.9<br>(42.5-51.0) | 47.9<br>(47.0-53.0) | 48.9<br>(46.3-53.5) | 44.6<br>(41.9-47.5) | 2.48 | .01 | 0.45 |
| Trait anxiety | 55.0<br>(51.0-58.6) | 47.0<br>(42.5-48.0) | 44.6<br>(43.3-44.8) | 52.9<br>(47.5-59.0) | 52.45<br>(47.0-56.7) | 52.0<br>(47.5-58.0) | 54.5<br>(48.0-59.8) | 49.6<br>(46.3-50.7) | 2.94 | .003 | 0.54 |
| Secondary outcome measures |  |  |  |  |  |  |  |  |  |  |  |
| ISI | 14.0<br>(12.0-14.5) | 9.8<br>(8.5-10.4) | 8.4<br>(8.2-8.7) | 12.0<br>(9.0-13.5) | 12.9<br>(9.0-14.5) | 8.5<br>(4.5-11.0) | 13.5<br>(10.5-14.8) | 9.1<br>(5.8-11.0) | 4.33 | <.001 | 0.79 |
| DERS-SF |  |  |  |  |  |  |  |  |  |  |  |
| Strategies | 9.0<br>(7.0-10.5) | 6.2<br>(5.0-6.2) | 8.1<br>(6.5-8.1) | 8.2<br>(6.0-10.0) | 9.0<br>(6.5-11.5) | 8.3<br>(7.5-9.0) | 9.0<br>(7.0-11.0) | 7.3<br>(6.0-7.8) | 3.22 | .001 | 0.59 |
| Non-acceptance | 8.3<br>(7.0-9.5) | 6.8<br>(5.5-6.9) | 8.1<br>(7.1-9.1) | 8.5<br>(6.5-10.5) | 7.7<br>(6.9-8.5) | 7.6<br>(6.0-8.5) | 8.0<br>(7.0-9.0) | 7.2<br>(6.0-7.8) | 0.84 | .41 | 0.15 |
| Impulse | 7.8<br>(5.5-9.0) | 5.2<br>(5.1-6.0) | 6.9<br>(6.0-6.9) | 7.5<br>(6.0-8.5) | 8.5<br>(6.0-9.5) | 7.4<br>(6.0-6.9) | 8.1<br>(6.0-9.0) | 6.4<br>(6.0-6.4) | 3.12 | .002 | 0.57 |
| Goals | 10.6<br>(9.0-12.0) | 8.6<br>(7.8-9.5) | 10.6<br>(10.0-10.6) | 10.3<br>(7.0-13.0) | 10.2<br>(8.0-12.5) | 10.8<br>(10.8-12.0) | 10.4<br>(9.0-12.0) | 9.7<br>(9.2-11.0) | 1.40 | .17 | 0.26 |
|  | Week 1 | Week 4 | Week 7 | Week 1 | Week 4 | Week 7 | Pre-intervention | Post-intervention |  |  |  |
| Awareness | 10.0<br>(9.0-12.0) | 10.6<br>(10.6-11.5) | 12.13 (12.1-13.0) | 10.0<br>(8.0-12.0) | 10.7<br>(9.0-12.5) | 10.4<br>(9.0-10.7) | 10.5<br>(9.0-12.8) | 10.5<br>(9.0-11.0) | 0.41 | .69 | 0.08 |
| Clarity | 7.8<br>(6.5-8.5) | 7.3<br>(6.0-7.3) | 7.4<br>(5.5-7.4) | 7.7<br>(6.5-9.5) | 6.4<br>(6.0-6.9) | 7.7<br>(7.0-8.0) | 7.1<br>(6.0-8.0) | 7.5<br>(6.0-7.9) | -0.58 | .57 | 0.11 |
| MAAS | 50.9<br>(47.0-54.0) | 61.1<br>(60.6-64.0) | 65.4<br>(64.7-66.7) | 47.4<br>(36.5-55.0) | 48.7<br>(44.5-57.0) | 57.5<br>(50.0-60.25) | 49.9<br>(46.3-54.0) | 59.2<br>(50.5-63.8) | -3.85 | <.001 | 0.70 |
| DASS-21 |  |  |  |  |  |  |  |  |  |  |  |
| Depression | 8.0<br>(7.0-9.6) | 5.3<br>(3.0-5.3) | 5.4<br>(5.0-5.4) | 10.3<br>(7.5-14.5) | 10.4<br>(9.2-14.0) | 7.4<br>(6.0-8.5) | 9.7<br>(7.0-13.3) | 6.4<br>(4.0-6.9) | 3.34 | <.001 | 0.61 |
| Anxiety | 8.0<br>(7.5-9.0) | 4.6<br>(3.5-5.8) | 3.6<br>(3.0-3.6) | 7.0<br>(4.0-8.5) | 6.7<br>(5.0-8.0) | 4.6<br>(2.0-5.5) | 7.7<br>(7.0-9.0) | 4.6<br>(3.0-5.8) | 3.43 | <.001 | 0.63 |
| Stress | 12.0<br>(10.0-14.0) | 7.6<br>(5.5-7.8) | 6.6<br>(6.6-7.0) | 11.0<br>(8.0-13.0) | 11.2<br>(5.0-5.4) | 10.3<br>(10.0-12.5) | 11.7<br>(10.0-12.8) | 9.0<br>(8.0-10.0) | 2.95 | .003 | 0.54 |

*Note.* INTG = intervention group; WCG = waitlist control group; IQR = interquartile range (25<sup>th</sup>-75<sup>th</sup> percentile); BAI-II = Beck Anxiety Inventory II; STAI = State-trait Anxiety Inventory-State; PSQI = Pittsburg Sleep Quality Index; ISI = Insomnia Severity Index; DERS-SF = Difficulties in Emotion Regulation Scale short-form; MAAS = Mindfulness Awareness Attention Scale, DASS-21 = Depression Anxiety Stress Scale 21-items

### Physiological outcomes

Per-protocol analyses were conducted for physiological data (*n*=18). Four participants were excluded from the analyses due to missing (*n*=2) or noisy (*n*=2) EEG data. The AF7 channel of two additional participants showed higher noise content and were also excluded from analyses. Pre- and post-intervention changes in EEG power feature distributions (Supplement 1) as well as the correlation between changes in mean EEG features and changes in primary outcomes (Supplement 2) for the remaining participants were analyzed.

As shown in Table 3, mean alpha-band power features indicated decreased arousal from pre- to post-intervention for most participants. Employing a statistical threshold of *p*<.05, 66.7% (8/12) exhibited a significant increase in post-intervention mean relative alpha power of AF7 compared to pre-intervention, and 71.4% (10/14) showed a significant increase in that of AF8. Fifty percent of participants exhibited a reduction in mean beta-2 relative power post-intervention compared to pre-intervention for AF7 (6/12) and AF8 (7/14), with 53.8% of these changes reaching statistical significance. Similarly, for beta-3 power, 50% for AF7 (6/12) and 71.4% for AF8 (10/14) showed reductions, with 81.3% being statistically significant. In contrast, fewer participants exhibited reductions in mean beta-1 power, being 50% for AF7 and 28.6% for AF8.

**Table 3.**
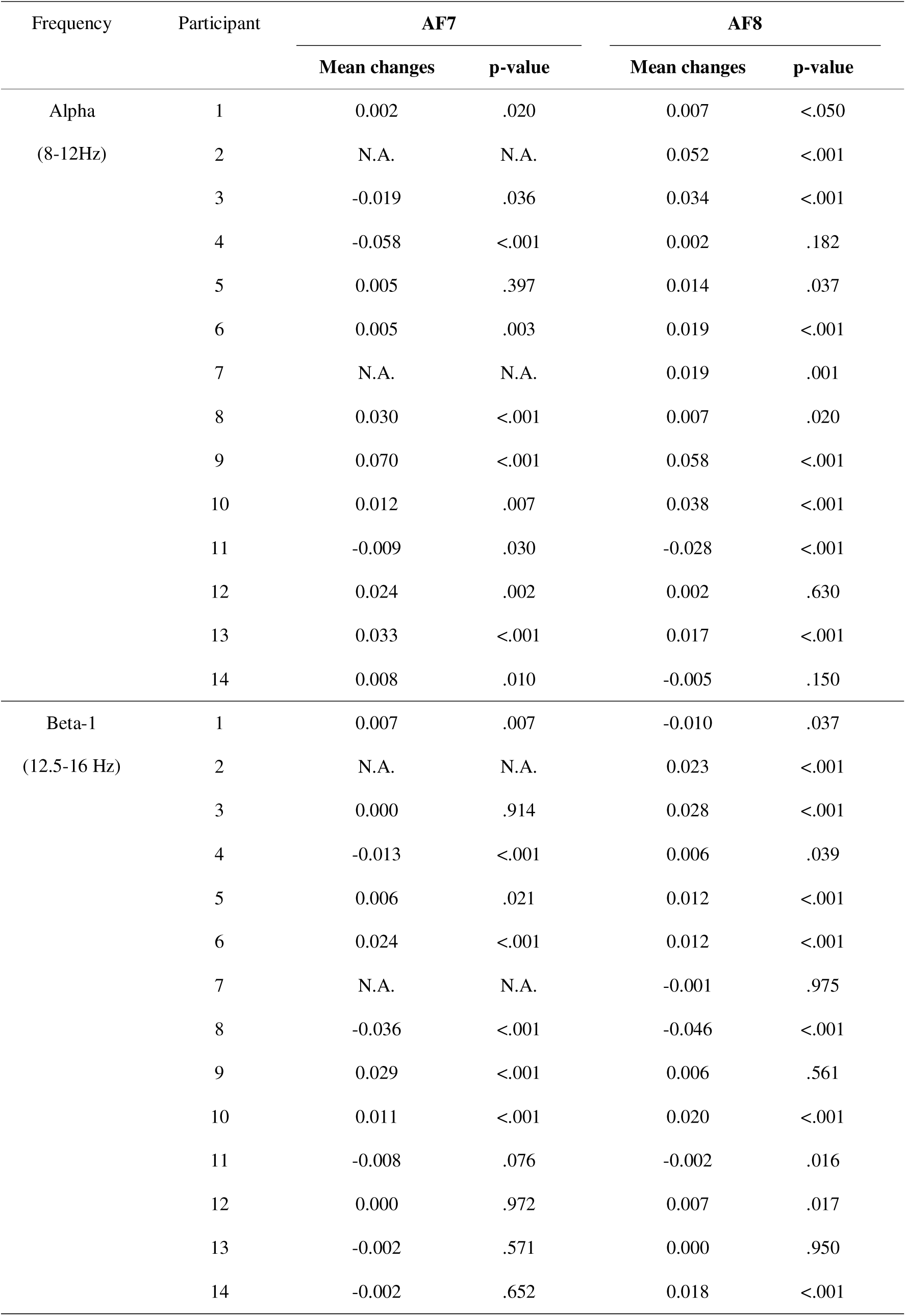
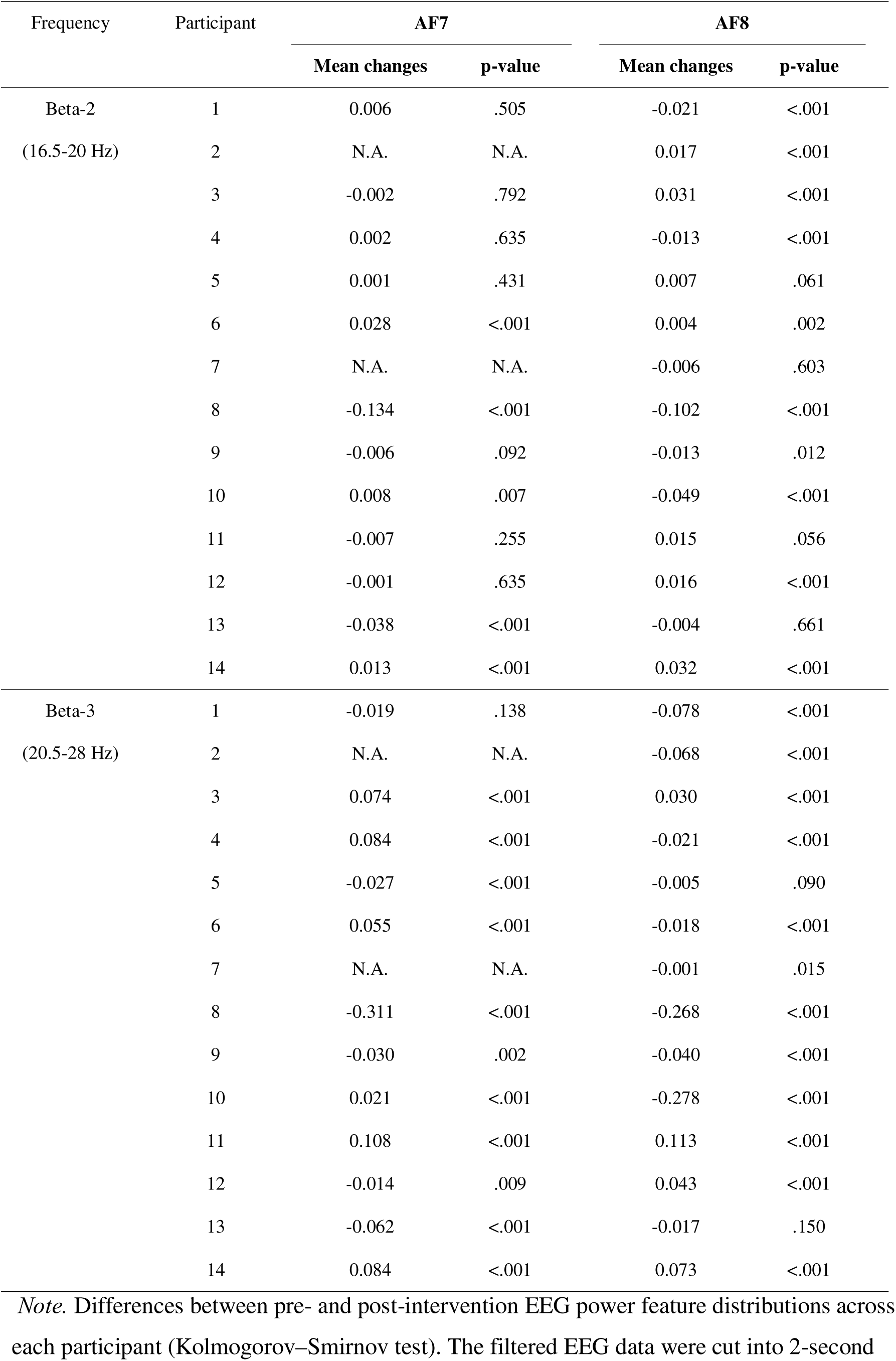
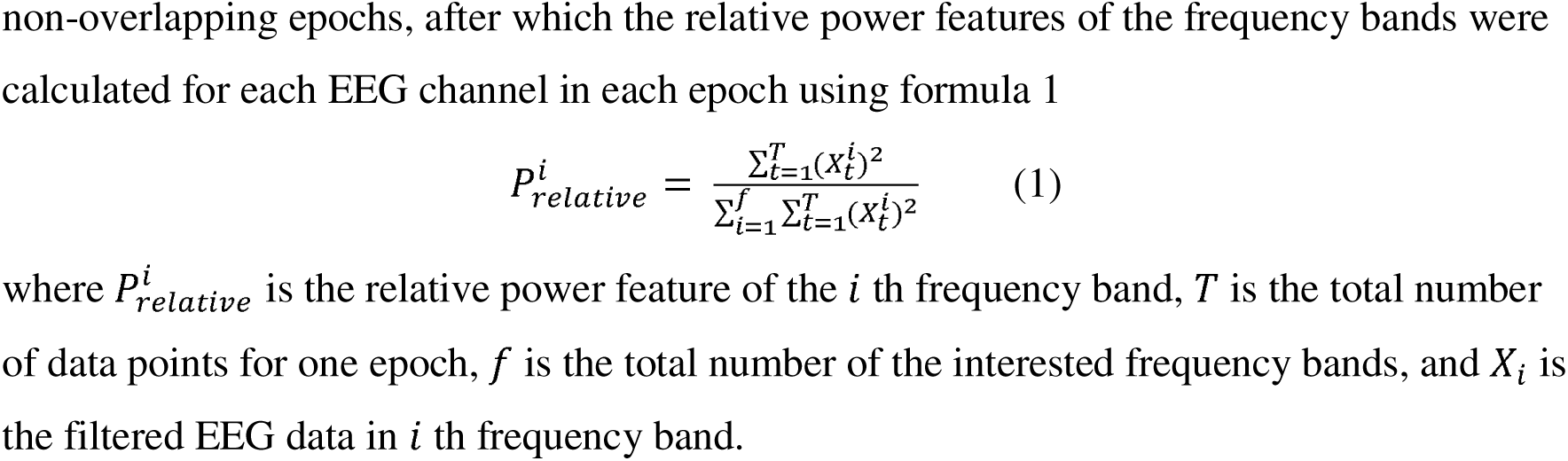
Physiological changes in AF7 and AF8 from pre- to post-intervention.

Supplement 2 illustrates the correlations between changes in mean power features and primary outcomes (STAI-S, STAI-T, BAI-II). There was a negative and marginally significant correlation between changes in mean alpha-band relative power of AF7 and STAI-S (*r*=-.53, *p*=.075). For the beta-2 band, significant positive correlations were observed between changes in mean relative power of AF7 and STAI-S (*r*=.65, *p*<.05), and AF8 and BAI-II (*r*=.58, *p*<.05). No other significant correlations were observed. The majority of alpha-band power feature changes negatively correlated with those of primary outcomes (83.3%), while a similar proportion of beta-band changes positively correlated with them (83.3%).

## Discussion

This study reports the safety, acceptability, and preliminary efficacy of a novel, personalized, BCI-based intervention to entrain mindful anxiety regulation among individuals with anxiety. Our intervention combines BCI technology, gamification, and mindfulness principles to deliver treatment that can be self-administered in the home setting. Overall, the findings of the present study are favorable.

While the intervention was safe, a third of the participants reported connectivity issues with the headset, which interrupted gameplay and contributed to study attrition. Some participants may have been under stress due to these technical difficulties, reflected in their physiological data. Although participants were taught how to use the headset, unfamiliarity with it renders difficulties in application possible. Despite this, qualitative feedback indicates that participants found the intervention helpful. The high treatment completion and adherence rates suggests that eight sessions of psychoeducation and BCI training over a 2-week period is acceptable to participants.

Further, the results indicated significant improvements in symptoms of anxiety, depression, and insomnia after two weeks of eight intervention sessions. Consistent with previous research [e.g., 27], the present study found evidence following the intervention for reduced anxiety that is associated with higher brain alpha activity and lower beta activity. However, mixed findings across beta-1, beta-2, and beta-3 waves were found, where some participants showed elevated activity post-intervention. One possible explanation for the elevation in beta activity is the increase in focus and concentration [29, 30]. Participants could be in a state of relaxation, with higher alpha and lower beta activity, or mindful awareness, associated with higher beta activity.

Indeed, findings indicated significantly greater mindful awareness at post-intervention, providing support that the intervention assisted in entraining mindfulness. Mindfulness develops over time and requires an ongoing commitment to practice [31]. The regular cycles of anxiety elevation and mindful relaxation during each session, as well as the consistent practice over two weeks, could be feasible for cultivating mindfulness in the short term. Future studies would need to examine the sustainability of these effects.

Participants demonstrated significantly lower difficulties with emotion regulation after the intervention. Specifically, participants reported significant increases in their ability to employ situationally appropriate emotion regulation strategies and to control impulsive behaviors when experiencing negative emotions. This finding concurs with existing studies that show the inverse relationship between mindfulness and emotion regulation difficulties [32, 33] and provides additional support for our intervention that entrains mindful anxiety regulation.

## Limitations and future research

There was decreased statistical power in the present study resulting from an attrition rate that was greater than anticipated (40%). Future research could include a larger sample size and refine the program to reduce the likelihood of technical difficulties. Future trials should also consider including double-blinding and an active control condition, such as a computerized training [34] to minimize response bias and rule out effects of simply participating in an online program.

## Conclusions

Our personalized, BCI-based, mindfulness intervention is a feasible and potentially efficacious treatment that can be self-administered at home. The promising findings warrant a larger clinical trial.

## Data Availability

All data produced in the present study are available upon reasonable request to the authors

## Abbreviations

BAI-II: Beck Anxiety Inventory II
CBT: cognitive behavior therapy
DASS-21: Depression Anxiety Stress Scale 21-items
DERS-SF: Difficulties in Emotion Regulation Scale short-form
EEG: electroencephalography
INTG: intervention group
ISI: Insomnia Severity Index
MAAS: Mindfulness Awareness Attention Scale
SSRIs: selective serotonin reuptake inhibitors
STAI: State-trait Anxiety Inventory
WCG: waitlist control group

