## Supplementary figures and images for "Personalized Brain-Computer Interface-based Intervention for Mindful Anxiety Regulation in Young Adults: A Randomized Clinical Trial"

### Supplement 1

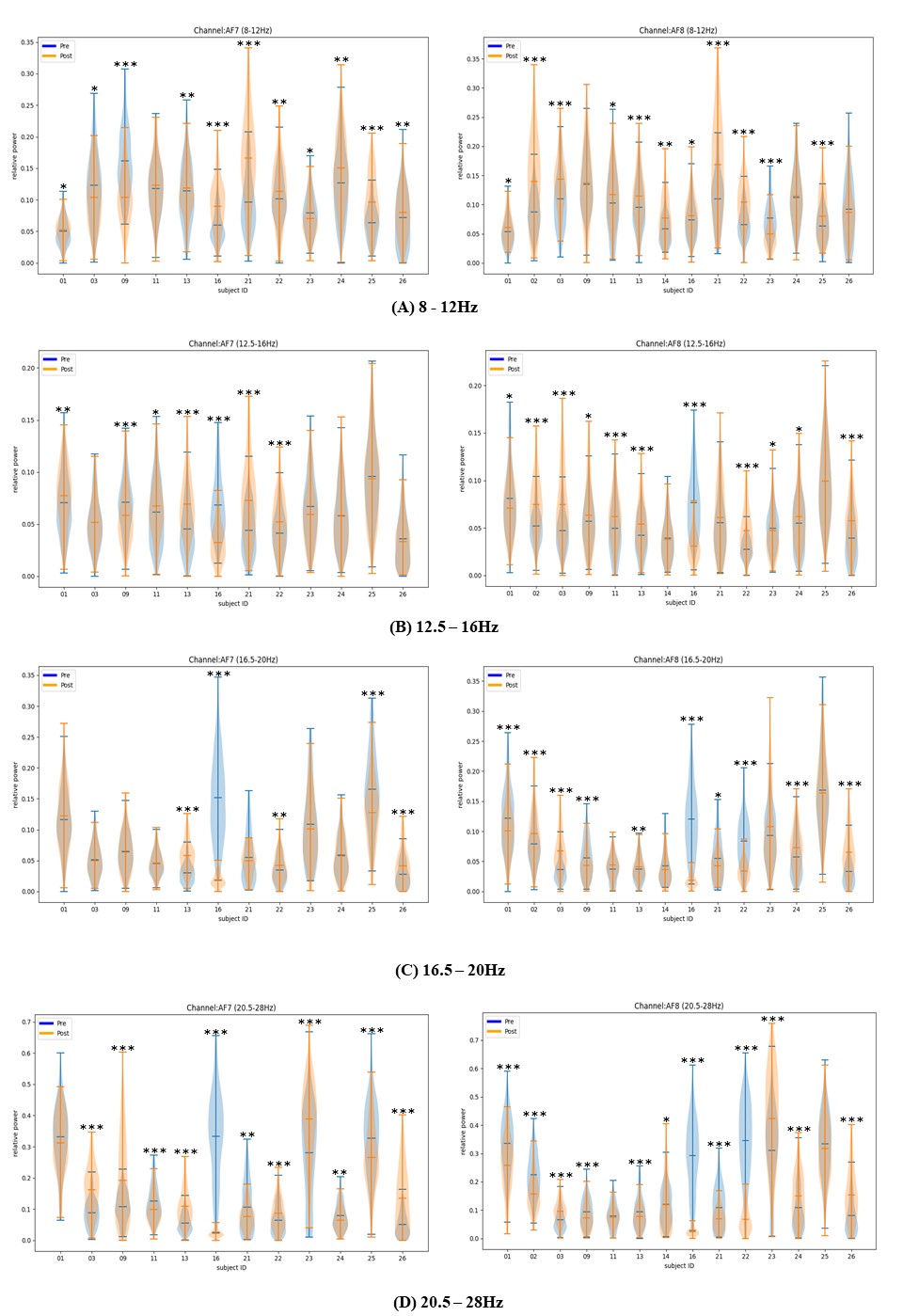

### Supplement 2

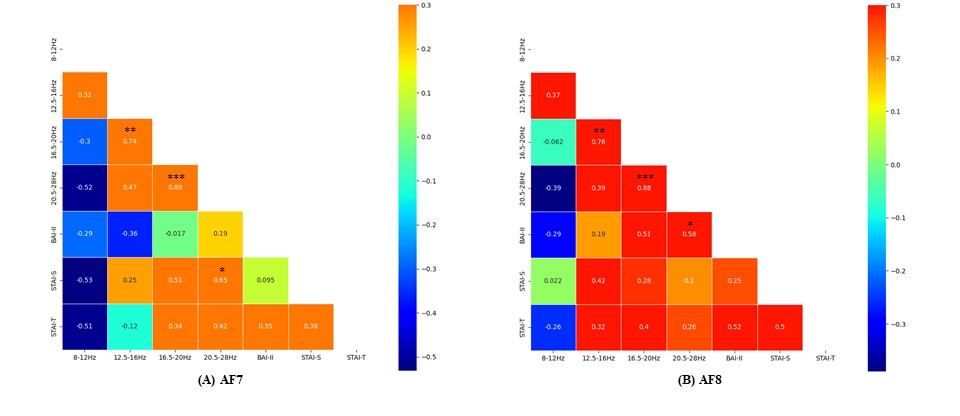
